# PALLIATIVE CARE INTEGRATED INTO PRIMARY HEALTHCARE SERVICE (PALLI – PHC): AN IMPLEMENTATION STUDY IN ROHINGYA REFUGEE SETTING IN COX’S BAZAR, BANGLADESH

**DOI:** 10.64898/2026.02.03.26345518

**Authors:** Charls Erik Halder, Syed Sunny Uz Zaman, Md Abeed Hasan, Md Mostafizur Rahman, Md Abu Sayum, Emmanuel Roba Soma, James Charles Okello

## Abstract

In humanitarian crises, health services focus on acute lifesaving care, and the needs of patients with chronic, progressive, and/or life-limiting illnesses are usually neglected. To address this gap, the International Organization for Migration (IOM) has been implementing an integrated palliative care service model: Palliative Care Integrated into Primary Healthcare (PALLI-PHC). This study aims to assess service outputs and patient-level outcomes of the model in the Rohingya refugee camps.

PALLI-PHC consists of a service delivery system supported by a health system support chain. The service is delivered at facility and home levels with an established referral pathway, triage and assessment, clinical evaluation, and care planning. The support chain comprises multidisciplinary engagement, health workforce, advocacy and capacity building, financing, community engagement, essential medical logistics, and information management.

Between January 2020 and December 2024, the programme delivered 40,776 consultations for a cohort of 11,599 patients. The top conditions managed were stroke, cancers, diabetic complications, hypertensive complications, and COPD. Prevalent symptoms included pain, fatigue, anxiety and worry, depressed mood, shortness of breath, and insomnia. Care was provided through facility-based and home-based services, including pain and symptom management, psychosocial and spiritual support, rehabilitation, caregiver support, and end-of-life care. Across follow-up visits, mean ESAS-r symptom scores declined significantly and the proportion receiving pain medication increased, although average disability scores also rose modestly over time.

This study shows how palliative care can be integrated into primary healthcare in a protracted humanitarian setting and can achieve measurable patient-level improvements, including reduced symptom burden and increased pain medication utilization.

## INTRODUCTION

Globally, patients living with serious health-related suffering in humanitarian settings remain largely neglected [1]. This burden is often intensified by acute crises, political instability and population displacement, where routine health services are disrupted and family support systems are fractured [2]. Palliative care seeks to relieve serious health-related suffering and support patients and families across physical, psychosocial and spiritual domains [1, 3–4].

The Rohingya refugee crisis in Cox’s Bazar, Bangladesh, illustrates this gap, where close to one million Rohingya refugees are living across 33 camps with limited access to specialist care and constrained resources [5]. Alongside high levels of trauma and acculturation stress, chronic illness and disability contribute to complex symptom burden and caregiver strain [6].

Refugee demographics extended across age groups from child to elder, and predominant disease demographics are non-communicable disease, tumors/cancers and organ failures with communicable disease outbreaks. PHC services includes maternal and child health, nutrition, sexual and reproductive health, communicable disease prevention/control, non-communicable disease treatment, and mental health and psychosocial support. Palliative care was deliberately integrated into this package as a cross-cutting service.

The PALLI-PHC model was developed by the International Organization for Migration (IOM) in response to these contextual needs, embedding palliative care within routine outpatient, inpatient and community-based primary healthcare services (see Supplementary Figure S1) [7]. The model aligns with WHO guidance for humanitarian emergencies and symptom relief and is informed by MSF clinical guidance adapted to resource-constrained settings [8–9].

Published experience on palliative care for displaced populations includes efforts to deliver symptom relief and supportive care through primary care teams and community outreach, but the evidence base remains limited [10]. A review of the Eastern Mediterranean Region also highlights substantial unmet palliative care needs among migrants and refugees, alongside limited service availability and policy capacity [11].

Despite growing global recognition of palliative care as part of universal health coverage and humanitarian response, evidence on sustained, integrated delivery models in protracted refugee settings remains limited [1, 8, 12]. Most published reports describe time-limited pilots or models that depend on specialist teams, with limited linkage to routine primary healthcare systems [8, 10, 12].

In response to this gap, the International Organization for Migration (IOM) implemented Palliative Care Integrated into Primary Healthcare (PALLI-PHC) in the Rohingya refugee camps and adjacent host communities in Cox’s Bazar, Bangladesh. Unlike stand-alone or specialist-dependent models, PALLI-PHC embeds palliative care within routine outpatient, inpatient, and community-based primary healthcare services, supported by structured workforce training, essential medicine integration, referral systems, and continuous monitoring. The objective of this study is to describe the PALLI-PHC model and assess its service outputs, patient characteristics, symptom burden patterns, and implementation performance in a protracted humanitarian setting.

## METHODOLOGY

### Study design

This was a retrospective observational implementation study assessing the design and performance of a palliative care model integrated into a primary healthcare system in a protracted refugee setting. The study analysed routinely collected, de-identified and anonymized programme data generated through service delivery to evaluate service coverage, patient characteristics, key palliative care conditions, symptom burden, and implementation challenges.

### Study setting

The study was conducted in the Rohingya refugee camps and adjacent host communities of Ukhiya and Teknaf, Cox’s Bazar, Bangladesh, one of the largest and most protracted humanitarian settings globally. Health services are delivered through a network of primary healthcare facilities supported by humanitarian partners under coordination mechanisms led by the health sector and the Government of Bangladesh. The palliative care programme was implemented by the International Organization for Migration (IOM) through selected primary healthcare facilities providing outpatient, inpatient, and community-based services under the PALLI-PHC model.

### Study population

The study population included patients with chronic and life-limiting illnesses who received palliative care services through the PALLI-PHC model during the study period. Both adult and paediatric patients were included. Patients were identified through routine clinical assessment and referral by community health volunteers and healthcare providers working in participating primary healthcare facilities.

### Data sources and data collection

De-identified programme data were extracted for the period January 2020 to December 2024. Data were originally recorded in facility registers and outpatient and inpatient reporting forms, entered electronically using Kobo Toolbox, and synchronised to a central database. Routine data quality checks, including verification and cleaning, were conducted as part of standard programme monitoring. Individual-level identifiers were not collected, accessed, or available to the study team at any stage. The anonymized dataset was accessed for research purposes between 01 August 2025 and 15 September 2025.

Variables included sociodemographic characteristics, primary diagnosis and comorbidities, services received, symptom burden (ESAS-r), functional status, and medicines prescribed.

### Data analysis

Data were analyzed using a combination of descriptive, regression, and trend analyses. Categorical variables were summarized using frequencies and percentages, while continuous variables were summarized using means or medians, as appropriate.

At the individual level, regression analyses were conducted to examine associations between patient characteristics, clinical conditions, and symptom burden and functional outcomes, using de-identified patient-level data. These analyses were intended to explore relationships between variables rather than to estimate causal effects.

At the population level, visit-level aggregation was used to describe programme-wide patterns in symptom burden, pain medication utilization, and functional status over time. Mean symptom scores and proportions were calculated across follow-up visits to characterize overall trends in service utilization and patient outcomes.

### Intervention

The International Organization for Migration (IOM) implemented the Palliative Care Integrated into Primary Healthcare (PALLI-PHC) model to integrate palliative care into routine primary healthcare services for Rohingya refugees and adjacent host communities in Cox’s Bazar, Bangladesh. The model has two linked parts: a service delivery system and a health system support chain that enables delivery and continuity (PALLI-PHC Model – Supplementary File S1).

During each consultation, symptoms were assessed using the Edmonton Symptom Assessment System Revised (ESAS-r), and scores were recorded in the patient chart [13–15]. Functional status was assessed using the Palliative Performance Scale v2 (PPSv2) [16]. Prognosis was estimated using the Palliative Prognostic Index (PPI), calculated from PPSv2 and clinical indicators [17]. Pain was assessed using age-appropriate tools: the FLACC scale for young children and the Faces Pain Scale-Revised for children and adolescents, alongside the numerical rating scale for adults [18–19]. Based on findings, an individualized care plan was developed in collaboration with patients and families.

Management followed stepwise approaches to analgesic selection and escalation consistent with WHO guidance, alongside non-pharmacological measures, psychosocial support and caregiver counselling as available [8, 20].

PALLI-PHC delivers holistic care by integrating mental health and psychosocial support, social support, spiritual care, rehabilitation, and end-of-life care. The team coordinates with facility-based psychologists and community volunteers for counseling and distress management, supports caregivers through education and practical guidance, and collaborates with religious leaders to provide culturally appropriate spiritual support. Rehabilitation services include physiotherapy and assistive devices to maximize comfort, mobility, and daily functioning. End-of-life care focuses on comfort, dignity, preference-based care planning, and bereavement support.

Client and family satisfaction was assessed using the FAMCARE scale [21].

## ETHICAL CONSIDERATION

The programme is part of IOM’s health response in the Rohingya refugee crisis being implemented in coordination with the Office of the Civil Surgeon and the Relief and Repatriation Commissioner (RRRC) Health Coordinator, who oversee health activities within the Rohingya refugee response in Cox’s Bazar, Bangladesh. Ethical clearance for this study was obtained from the Institutional Review Board/Ethics Review Committee (IRB/ERC) of North South University (Approval No. 2025/OR-NSU/IRB/0510) for retrospective programme data use. Internal approval was also obtained from the International Organization for Migration (IOM) Migration Health Research Unit both for the study and retrospective programme data use for the study.

This study was a retrospective observational analysis using routinely collected programme data generated through service delivery under the PALLI-PHC model. The data analyzed were collected as part of routine public health response and health service delivery activities between January 2020 and December 2024 and were subsequently used for research purposes under an approved secondary-use protocol. Original clinical records were accessed for research purposes between 01 August 2025 and 15 September 2025.

At no point did the investigators have access to direct or indirect patient identifiers. All individual-level data were de-identified and anonymized prior to analysis, and findings are reported in aggregate form only. As the data were collected as part of routine clinical care and programme monitoring, no additional data were collected from patients for research purposes. Informed consent was obtained from patients or the care givers for the use of their information for monitoring, evaluation and research.

## RESULTS

Between January 2020 and December 2024, IOM conducted 40,776 palliative care consultations (service contacts) among 11,599 patients. Data presented in this section were routinely collected as part of service delivery between January 2020 and December 2024 and analyzed retrospectively. This paper’s analysis is based on the number of palliative care consultations.

### Demographic profile of the patients

Majority of the clients accessed palliative care belonged to the age groups between 45 – 59 years (30.96%) and 60 – 74 years (36.08%). Both male and female patients accessed the care almost at equal proportions. Three-fourths of the patients were from the refugee community. Most were illiterate (55.19%) or could only sign (40.34%). The demographic characteristics of patients are shown in Table 1.

**Table 1:** Demographic profile of the patients (N=40776).

| Demographic Variable | Number | Percentage |
| --- | --- | --- |
| **AGE** |  |  |
| 0-17 years | 970 | 2.38% |
| 18-29 years | 1450 | 3.56% |
| 30-44 years | 6188 | 15.18% |
| 45-59 years | 12623 | 30.96% |
| 60-74 years | 14713 | 36.08% |
| 75+ years | 4832 | 11.85% |
| **GENDER** |  |  |
| Male | 20637 | 50.61% |
| Female | 20139 | 49.39% |
| **STATUS** |  |  |
| Host | 9975 | 24.46% |
| Refugee | 30801 | 75.54% |
| **Education** |  |  |
| **No formal education** | 22505 | 55.19% |
| Only can sign | 16450 | 40.34% |
| Primary (1-5) | 1216 | 2.98% |
| Secondary (6-10) | 498 | 1.22% |
| Degree | 84 | 0.21% |
| **Marital status** |  |  |
| Divorced | 25 | 0.06% |
| Married | 36703 | 90.01% |
| Separated | 59 | 0.14% |
| Unmarried | 1128 | 2.76% |
| Widowed/widower | 2861 | 7.01% |
| **Religion** |  |  |
| Buddhist | 20 | 0.05% |
| Christianity | 1 | 0.002% |
| Hindu | 124 | 0.30% |
| Islam | 40631 | 99.64% |
| **Occupation** |  |  |
| Jobless | 27136 | 66.55% |
| Housewife | 11599 | 28.45% |
| Day labor | 1672 | 4.10% |
| Farmer | 70 | 0.17% |
| Others | 299 | 0.73% |

### Key Conditions

The major conditions among patients receiving palliative care are illustrated in Figure 2a. The most prevalent diagnosis was stroke (37.7%), followed by carcinoma (10.5%), complicated diabetes (8.3%), hypertensive complications (8.2%), chronic abdominal disorders (7.3%), and chronic obstructive pulmonary disease (COPD) (7.0%). Other diagnoses included chronic liver disease (CLD) (4.3%) and additional chronic respiratory conditions. Ischaemic heart disease (IHD) (2.9%) and chronic kidney disease (CKD) (1.6%) were also identified but accounted for a smaller proportion of the total case mix.

**Figure 1:**
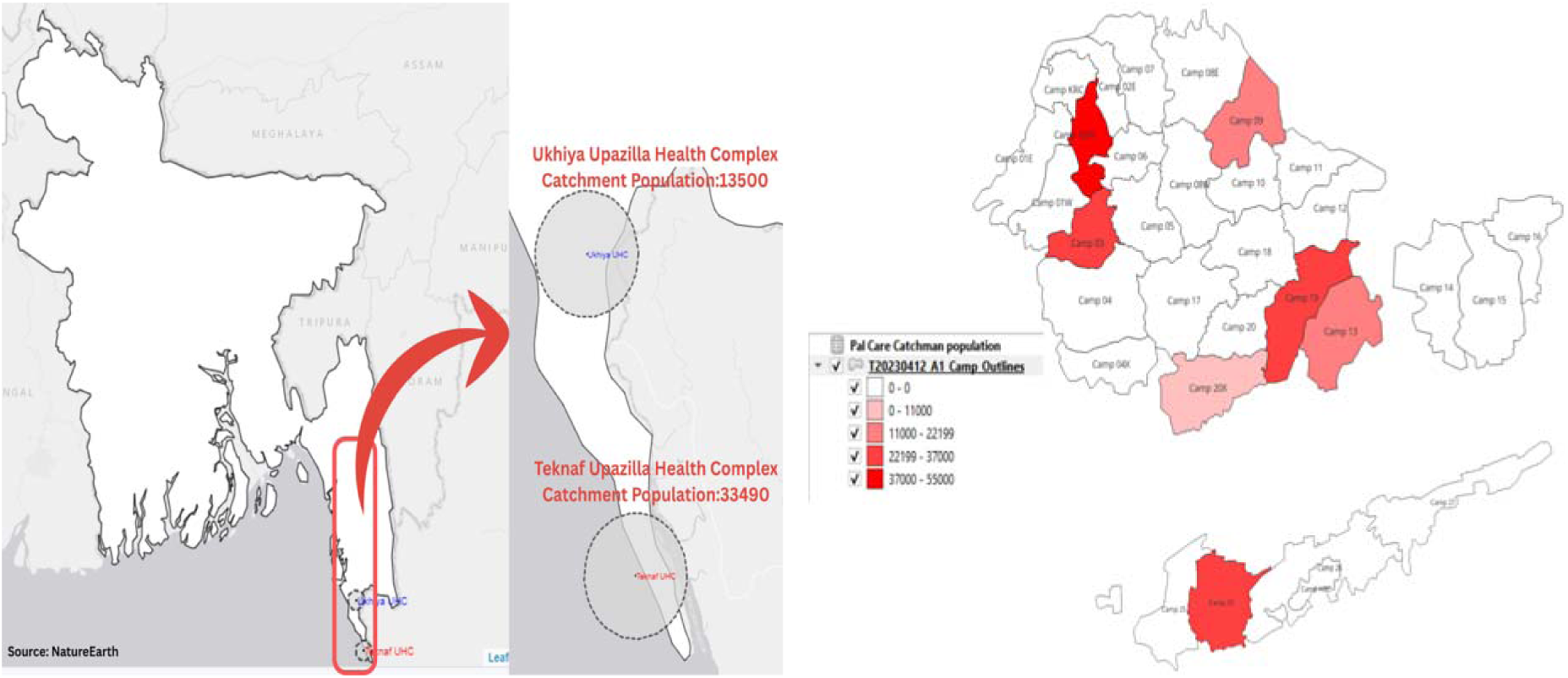
Palliative care catchment area with target population density in camps and in host.

**Figure 2:**
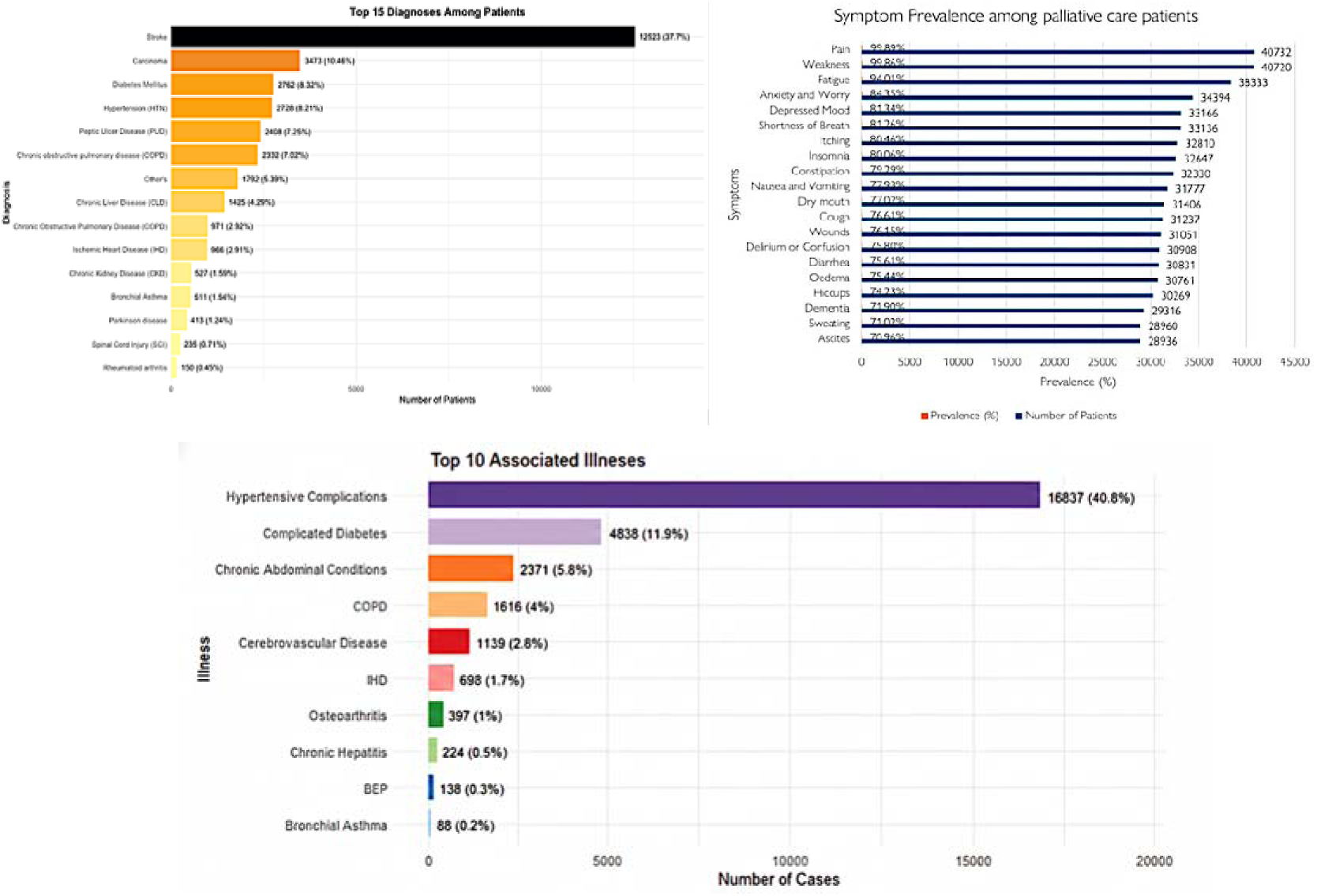
Top diagnoses along with symptoms prevalence and associated illness.

Figure 2b summarizes symptom burden among palliative care patients and shows a clear clustering of symptoms by prevalence. A high-burden symptom group was dominated by weakness (∼90%), pain (∼75%), and fatigue (∼74%), followed by psychological distress, including anxiety and worry (∼61%), depressed mood (∼47%), and insomnia (∼42%). A moderate-burden group (approximately 10%–35%) included itching, dry mouth, delirium/confusion, constipation, dementia, shortness of breath, cough, and nausea/vomiting, indicating a substantial proportion of patients experienced additional distressing symptoms beyond pain and fatigue. A lower-burden group (<10%) included wounds, oedema, sweating, loss of appetite, reduced well-being, ascites, hiccups, and diarrhoea.

Figure 2c illustrates the top ten comorbid illnesses among palliative care patients, with hypertensive complications (40.8%) being the most prevalent, followed by complicated diabetes (11.9%) and chronic abdominal conditions (5.8%). Other notable comorbidities included COPD (4.0%), cerebrovascular disease (2.8%), and ischaemic heart disease (1.7%).

### Type of intervention

The programme provided both a) conventional treatment support including palliative care medicines (92%) and curative care (chemotherapy, radiotherapy, surgery); and b) complementary care including physiotherapy (92.8%), social care (56%), psychological support (40%), spiritual support (10%) and traditional medicine (3%). The distribution of these treatment modalities is illustrated in figure 3.

**Figure 3:**
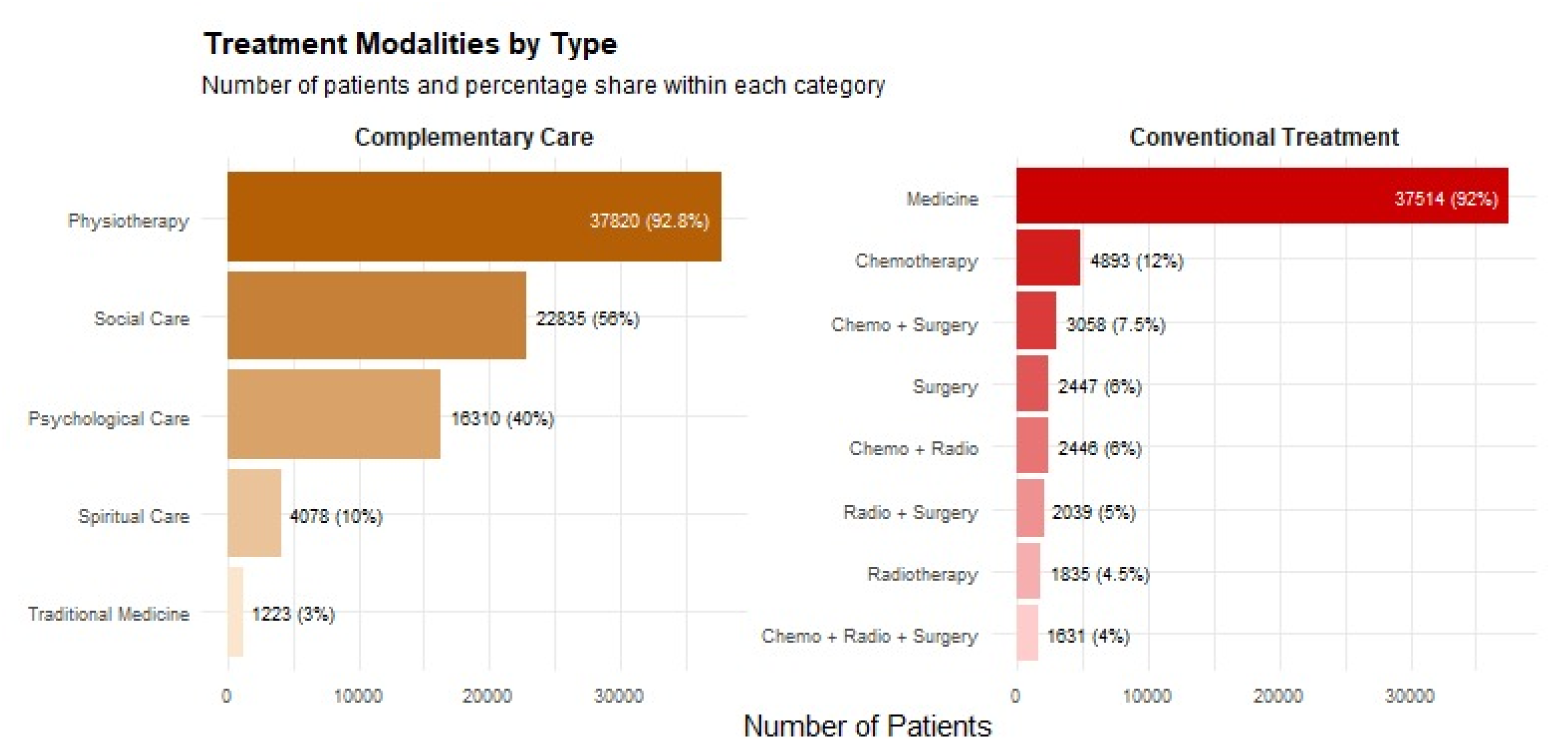
Treatment modalities in Palliative Care.

### Symptom burden

ESAS-r Total Score (0–100): Figure 4a demonstrates a trend of decreasing average symptom scores across the first seven follow-up visits. There was a clear drop in ESAS-r scores, particularly between the first and second visits, followed by minor fluctuations. This pattern reflects the effectiveness of palliative care follow-up in reducing symptom burden and enhancing comfort. Figure 4b demonstrates a trend of decreasing ESAS-r (100) scores with successive follow-up visits up to 7th visits followed by fluctuations indicating worsening symptoms in late follow-ups. A linear regression analysis confirms a statistically significant downward trend in symptom severity with each additional follow-up (β = –0.411, p < 0.001, R² = 0.007).

**Figure 4.**
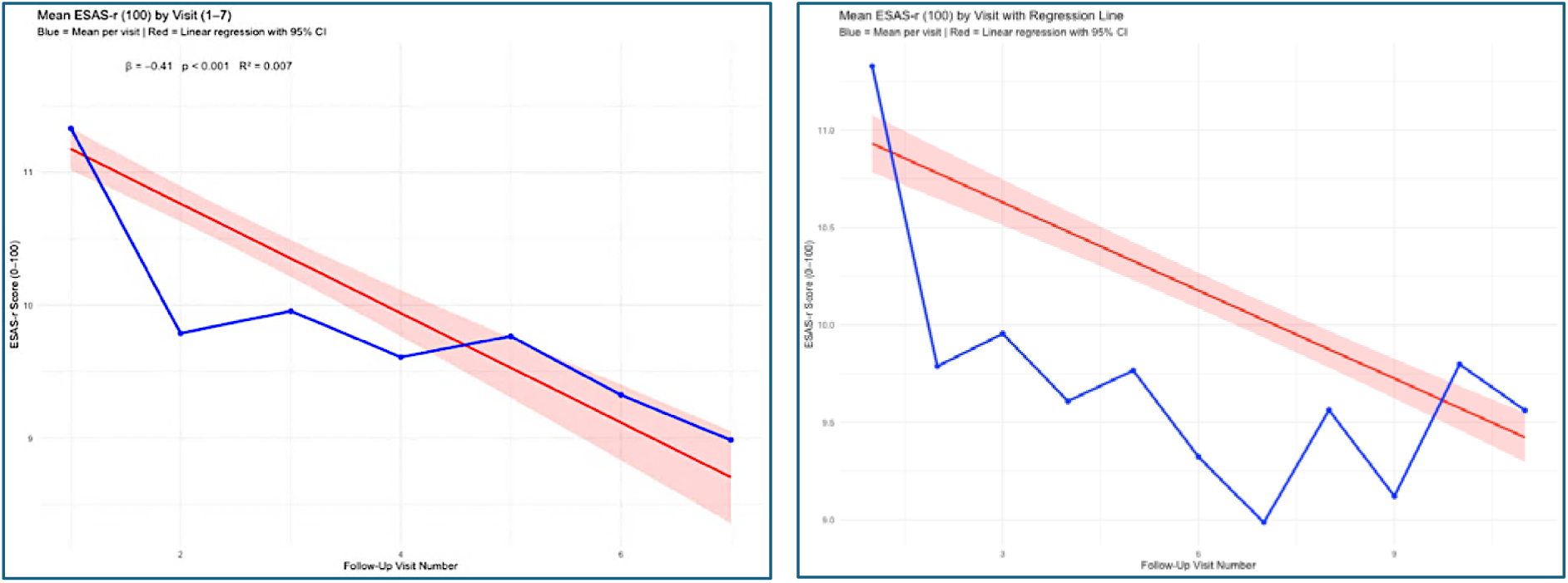
ESAS-r (Visits 1-7), ESAS-100 (includes all visits).

Importantly, the early and sustained decline through multiple visits suggests that symptom assessment and targeted management are producing measurable relief for most patients, while the late-visit fluctuations likely reflect a smaller, clinically more severe subgroup and disease progression rather than failure of care—highlighting the value of continued follow-up to identify deterioration early and adjust care plans accordingly.

### Pain Medication by Follow-Up Visit

Figure 5 shows a progressive increase in the proportion of patients receiving pain medication across follow-up visits. The blue line represents observed mean proportions at each visit, while the red line shows the fitted linear regression trend. Linear regression demonstrated a statistically significant upward trend in pain medication use over time (β = 0.013 per visit, p < 0.001, R² = 0.58). Although the increase per visit was modest, the overall pattern indicates progressively improved access to pharmacological pain management as follow-up continued. This trend is consistent with the program’s aim to systematically integrate pain management into routine palliative care services.

**Figure 5.**
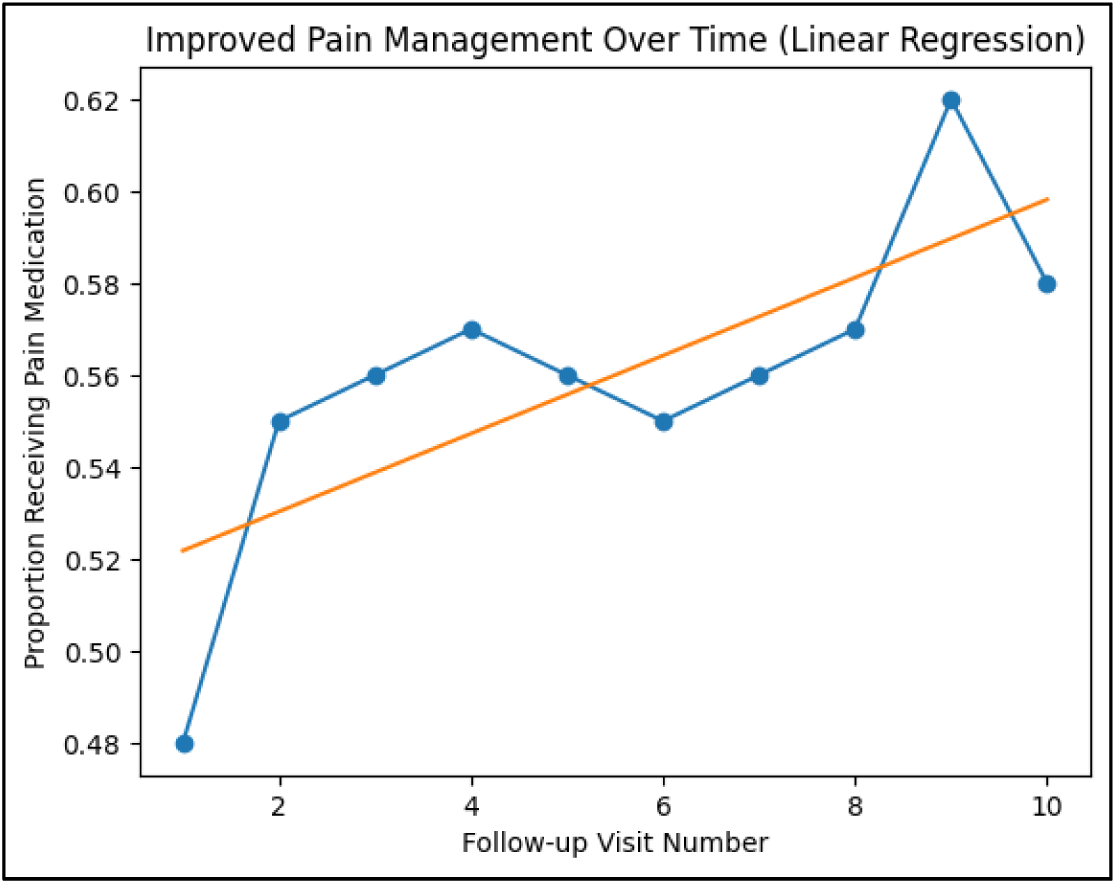
Pain Medication Over Time.

### Disability Score Regression Analysis

Supplementary Figure S2. Washington Disability Scoring indicates a mild upward trend in disability scores across all follow-up visits. While variability exists between visits, the overall average shows a statistically significant increase over time (β = 0.065, p < 0.001, R² = 0.011). This might reflect the progression of underlying conditions despite supportive care or a shift in focus from symptom relief to overall functionality. It reinforces the need for integrated rehabilitation services alongside palliative care, particularly for patients with long-term chronic conditions.

## DISCUSSION

To our knowledge, this is one of the first programme reviews to describe an integrated palliative care model embedded in routine primary healthcare delivery in a protracted refugee setting. Earlier work in the Rohingya context documented profound unmet palliative care needs and major gaps in access to basic symptom relief and supportive care [22]. Our findings show that a structured PHC-integrated model can deliver high service volume while addressing pain and other symptoms in a highly resource-constrained environment, consistent with global calls to reduce serious health-related suffering [1, 4, 8].

The Palliative Care Integrated into Primary Healthcare (PALLI-PHC) programme addressed these challenges by delivering facility- and home-based palliative care through trained multidisciplinary teams, standardized assessment and pain-management tools, reliable access to essential medicines, strong community and caregiver engagement, and coordinated referral, training, and monitoring systems embedded within existing primary healthcare services. The scale of service utilization—over 10,000 patients with chronic and life-limiting illnesses and more than forty thousand consultations—demonstrates both substantial unmet need and operational feasibility of integrating palliative care into routine PHC delivery in this setting.

The programme also served children and adolescents with serious illness and disability. Global guidance for humanitarian settings emphasizes that palliative care should be available across the life course, including for children [8, 12]. Many families faced low health literacy and strong stigma around severe illness. Clinician communication training, including the SPIKES framework and principles for serious-illness conversations, supported more sensitive delivery of difficult information [23–24].

Our caseload was dominated by stroke, cancers and complications of diabetes and hypertension, reflecting the growing burden of non-communicable diseases in humanitarian settings [5, 12]. This differs from cancer-centric palliative care patterns seen in many high-income settings and underscores the need for broad symptom relief and supportive services beyond oncology [1, 8, 12].

Fatigue emerged as a leading symptom across diagnoses, consistent with the multidimensional symptom burden captured by ESAS-based tools in palliative care populations [13–15]. Persistent fatigue may reflect advanced disease, deconditioning, anxiety and limited access to rehabilitation or nutritional support in camp environments.

Pain control remained a core challenge. Although use of pain medication increased across follow-up visits, opioid availability and prescribing remain constrained in many low-resource and humanitarian contexts [20, 25]. WHO guidance emphasizes stepwise analgesic escalation and access to essential medicines as foundational components of palliative care [4, 20, 25].

Psychological symptoms, including anxiety and depressed mood, were common. Mental health and psychosocial support is recommended as a core component of emergency response, and should be integrated with symptom management and caregiver support where possible [6, 8].

Rehabilitation needs were substantial, particularly among patients with stroke-related disability. Integrating basic physiotherapy, mobility aids and caregiver training may help preserve function and reduce caregiver burden in this setting.

Overall, the PALLI-PHC model demonstrates that palliative care can be embedded within routine primary healthcare services in a protracted humanitarian setting. The approach may help reduce serious health-related suffering while strengthening continuity of care for patients and families [1, 3–4, 7].

However, the model may not be directly generalizable to all humanitarian settings due to differences in workforce capacity, referral access and national policies related to opioid availability and essential medicines [20, 25].

## CONCLUSION

The PALLI-PHC model demonstrates that palliative care can be integrated into primary health care (PHC) in humanitarian crises in a practical and sustainable way, where people living with chronic and life-limiting illnesses face extreme vulnerability due to poverty, psychosocial stressors, and limited health system capacity. Despite major operational constraints, IOM implemented Palliative Care Integrated into Primary Healthcare (PALLI-PHC) in the Rohingya refugee camps and adjacent host communities, enabling structured symptom assessment, continuity of care, and supportive follow-up to reduce suffering and improve comfort.

This implementation experience shows that palliative care is needed for a broad spectrum of conditions in protracted crisis settings, including stroke, cancers, complications of non-communicable diseases, and chronic organ failure, alongside neurological, spinal, and immunological disorders. The high burden of pain, fatigue, weakness, psychological distress, and other symptoms highlights the necessity of a holistic approach that goes beyond cure-focused care. The findings support that with focused training for general health workers, task-sharing, standardized tools, and integration into existing PHC systems, palliative care can be delivered meaningfully even in resource-constrained humanitarian contexts. Future scale-up should prioritize expanding coverage through multi-partner adoption, strengthening rehabilitation and psychosocial components, and improving access to essential palliative medicines, including opioids, through coordinated policy and supply-chain solutions.

## Data Availability

The datasets generated and analysed during the current study are not publicly available due to ethical and data protection restrictions related to the study population but are available from the corresponding author upon reasonable request.

## ACKNOWLEDGEMENTS

CEH led, conceptualized and designed the study, provided overall methodological guidance, and led the drafting and critical revision of the manuscript. SSUZ and MAH assisted in refining the study design and drafting the manuscript. SUZ, MMR supported field coordination, MAH, MAS assisted supporting the data collection processes. JCO and ERS contributed to the critical review and refinement of the manuscript. JCO and ERS provided strategic oversight and ensured alignment of the study within broader programmatic and policy frameworks. All authors reviewed and approved the final version of the manuscript. CEH is the corresponding author and JCO is the guarantor of the study.

## GENERATIVE AI DISCLOSURE

Language editing assistance was provided using Grammarly (version 6.8.263) and ChatGPT (OpenAI; GPT-5 Thinking, accessed [November 2025]), limited strictly to grammar and clarity. No text, analyses, figures, or references were generated de novo. All authors reviewed and took full responsibility for the content.

---

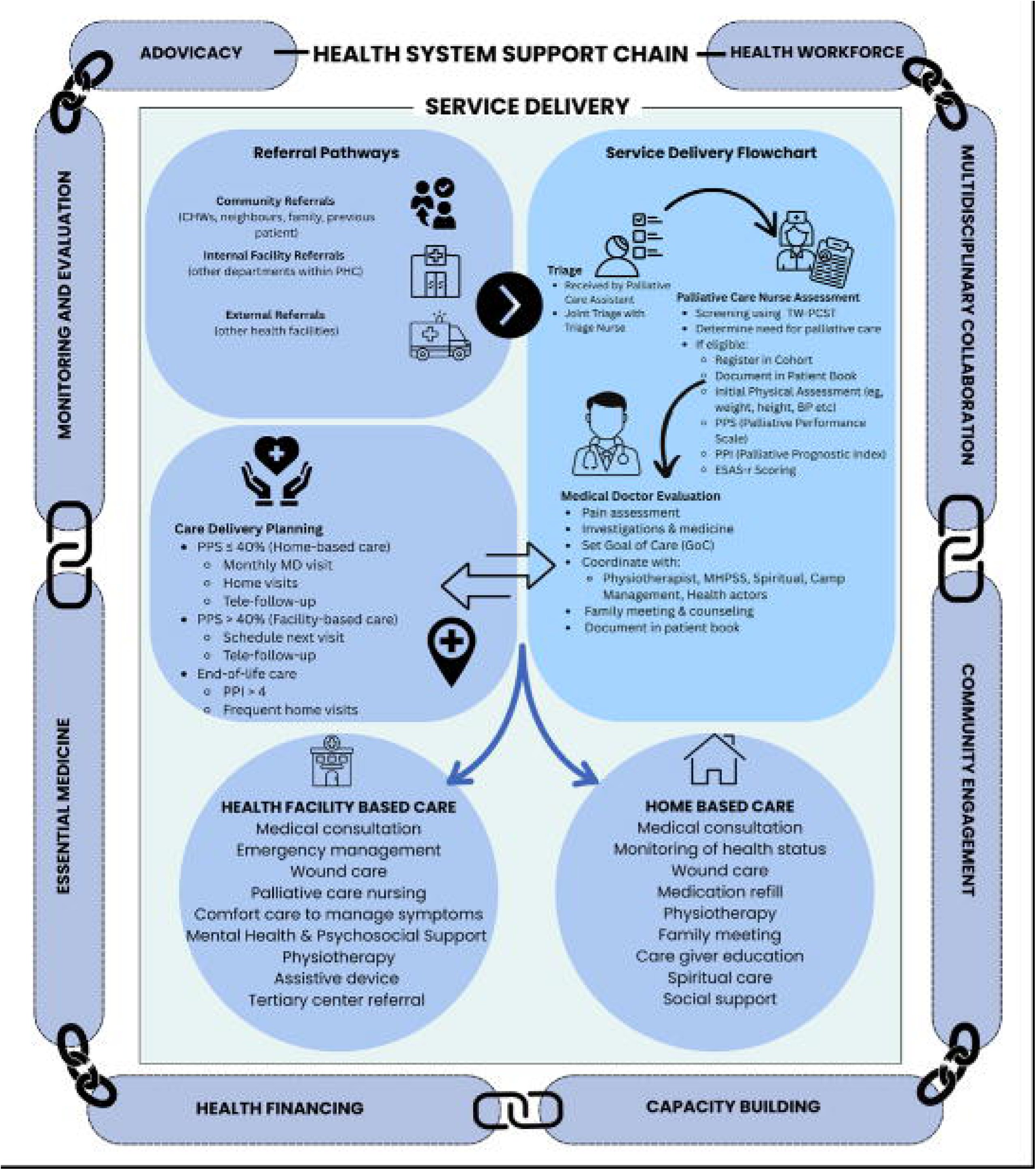

