## Supplementary File S1 for "PALLIATIVE CARE INTEGRATED INTO PRIMARY HEALTHCARE SERVICE (PALLI – PHC): AN IMPLEMENTATION STUDY IN ROHINGYA REFUGEE SETTING IN COX’S BAZAR, BANGLADESH"

**The PALLI-PHC Model**

IOM is implementing a Palliative Care Integrated into Primary Healthcare (PALLI-PHC) service model to integrate palliative care into the essential health service package for the Rohingya refugees in Cox’s Bazar. This service model comprises a service delivery system supported by a health system support chain (Supplementary Figure 1: PALLI-PHC Model).

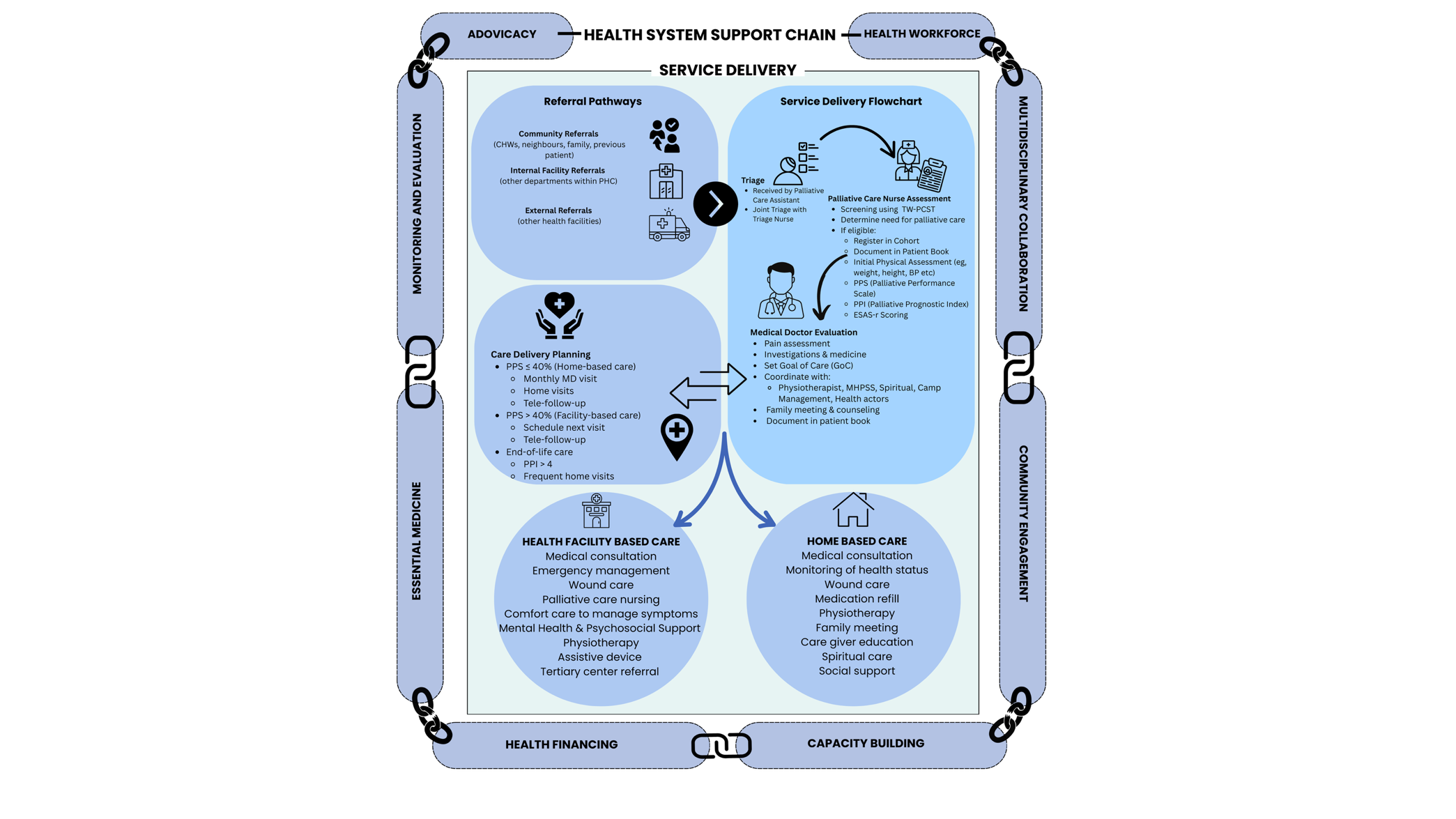

***Supplementary Figure 1: PALLI-PHC (Palliative Care Integrated into Primary Healthcare) Model***

The service is delivered at facility and home levels. The support chain enables effective integration and delivery of palliative care in primary healthcare settings. The PALLI-PHC model, aligned with WHO guidance, combines structured service delivery, capacity building, and outcome monitoring to provide holistic palliative care in crisis-affected primary healthcare. It is a feasible and replicable approach within other resource-limited refugee communities due to its implementation of capacity building, essential medicine, and community engagement components.

**SERVICE DELIVERY SYSTEM**

Under the PALLI-PHC model, IOM integrated palliative care into the essential health service package of its operated primary healthcare centers (PHCs). IOM palliative care includes health facility-based services, home-based care and community engagement. At the outpatient department, a trained palliative care team comprising a medical doctor, nurse, medical assistant, and palliative care assistants is deployed. The team also visits the inpatient department (IPD) to provide palliative support to the bed-ridden patients working closely with the IPD medical staff. The team offers home-based care to those who are non-ambulatory and unable to attend a clinic or wish for their end-of-life care at home. Palliative care assistants act as front-line workers to engage with the patient, families, community, and social and religious leaders.

Palliative care assessment: Patients requiring palliative care are referred to the palliative care team through various channels, including community health workers, clinicians, and/or triage at health facilities. All new patients undergo a series of assessments by the palliative care nurse using standard tools (Table 1) to plan holistic patient-centered care. This assessment enables the team to screen individuals requiring palliative care, measure symptom burden and functionality, and estimate the prognosis of patients [8] and thus, plan the modality of service, goal of care, and end-of-life care. The tools also allow follow-up of the improvement or deterioration of symptoms and functionality through the course of care.

**Supplementary Table 1: Palliative care patient assessment tools**

| Tool | Objective | Method | Interpretation | Frequency |
| --- | --- | --- | --- | --- |
| Palliative Care Screening Tool [9]  (Taiwanese Version-Palliative Care Screening Tool - TW-PCST) | Identify/screen patients requiring palliative care | Scores based on basic disease process, concomitant disease process, patient's functional status, and other symptoms. | Action recommended based on total score:   - 2: Give patient information about palliative care - 3: Consider palliative care consultation - 4: Requires palliative care | During registration |
| Revised Edmonton Symptom Assessment Scale (ESAS-r). [10] | Provide quantitative profile of symptoms over time | Patient rate intensity of each of the 9 common symptoms on a scale of 0 to 10 | For each symptom, 0 represents “no symptom,” and 10 represents “worst possible symptom.”  Allows monitoring of symptom intensity with the course of intervention | Each visit or daily (in IPD) |
| Palliative Performance Scale (PPS V2) [11] | To assess, document and monitor functional status | Measure the functional level of – a) ambulation, b) activities and extent of disease, c) self-care, d) food and fluid intake, and e) consciousness. | Measured at 10% decrements, where 100% indicates fully ambulatory and healthy, and 0% means the patient has died. | Each visit or daily (in IPD) |
| Palliative Care Prognostic Index (PPI) [11] | Estimate the prognosis of patients receiving palliative care, enabling them to set meaningful goals, priorities, and expectations of care | Score based on PPS, oral intake, oedema, dyspnea at rest, and delirium with a total ranging from 0 to 15 | >6 indicates survival of less than 3 weeks  >4 indicates survival of less than 6 weeks.  Determines the need for end-of-life care. | Each visit or daily (in IPD) |

**Goal of care planning:** In the first consultation, the palliative care team works with the patients and their care givers, and coordinates with different service points to set the goal of care so that the patients and/or their families can understand, adopt, and prepare for their upcoming necessary interventions.

**Pain and Symptom Management:** After initial screening and assessment by the triage nurse, the palliative care physician conducts detailed discussions with the patients and their families and plans the management of each symptom, aiming to address their physical, emotional, psychological, and spiritual needs [3].

Based on MSF and WHO guidelines, the programme uses the FLACC scale, Faces Pain Scale-Revised, and Numeric Rating Scale to assess pain severity across age groups [12–14].

**Supplementary Table 2: Pain assessment tool kit**

| Tool | Criteria | Method | Interpretation |
| --- | --- | --- | --- |
| FLACC Scale | Children <3 Years, patients with decreased consciousness levels or those unable to self-report | Score between 0 to 2 for each of 5 behaviors (Face, Legs, Activity, Cry, Consolability) | Total 0–10: 0 = Relaxed, 1–3 = Mild, 4–6 = Moderate, 7–10 = Severe pain |
| The Faces Pain Scale – Revised (FPS-R) | Children ≥3 Years or cognitively impaired adults | Patient points to a face that best describes their pain | Faces are scored as 0, 2, 4, 6, 8, 10.  0 = No pain  2–4 = Mild pain  6 = Moderate pain  8–10 = Severe pain |
| Numeric Rating Scale | Patients are able to self-report, usually adults. | Patient rates pain on a scale of 0 to 10 | 0 = No pain, 1–3 = Mild, 4–6 = Moderate, 7–10 = Severe pain |

The PALLI-PHC encourages several non-pharmacological and integrative approaches along with the analgesics. This includes heat/cold therapy, massage, relaxation, breathing techniques, music, meditation, and local herbs. The use of analgesics, especially Paracetamol, NSAIDs, tramadol, and oral morphine, as well as other adjuvants (e.g., antidepressants), is determined based on the pain scale using the WHO Analgesic Ladder [15]. One of the programme's unique innovations is integrating physiotherapy into a palliative care package, which provides additional pain management through manual and electro-therapy.

The palliative care team assesses and manages all other symptoms in coordination with clinical teams at PHCs, using pharmacological and non-pharmacological approaches as suggested in the standard guidelines [14, 16].

**Procedures and surgeries:** Basic procedures like paracentesis and pleural fluid aspirations are performed at the PHC level to relieve breathlessness resulting from ascites or pleural effusions [17]. Case by case, patients requiring advanced palliative surgery are referred to a government tertiary health facility at the Division level.

**Emergencies in palliative care**: The palliative care team tackles the common palliative care-related emergencies such as bleeding, agitation, convulsion, acute retention of urine, pleural effusion, massive ascites, etc. working closely with the emergency room teams at PHCs.

**Communication:** PALLI-PHC considers active listening as part of therapy and care. Therefore, the clinicians are expected to give a patient 5 to 10 times more time than a general physician [18]. Active listening strengthens the trust between patients and professionals. The " SPIKES " approach is used to deliver bad news to patients and/or family members [19].

**Mental Health and Psychosocial Support:** The palliative care team coordinates with the facility-based MHPSS team/psychologists for psychological support for patients and their families, which includes counseling, anxiety and depression management, cognitive behavioral therapy, mindfulness and relaxation, grief and bereavement support, and promoting family/caregiver support. The palliative care team also works with community-based MHPSS volunteers to provide psychological support at household level.

**Social support:** Social care is also a key component that includes but is not limited to meeting with and supporting the family members, providing them with the necessary information and emotional support, and training them to take care of the patients at home (e.g., nursing, dressing). Home-based palliative care team explores the social impact of illnesses and works closely with the family members and community to identify and address associated stigma and discrimination [1].

**Spiritual care:** PALLI-PHC considers spiritual care as part of compassionate and holistic care, acknowledging the need for spiritual well-being for those with life-threatening illnesses. Palliative care staff train and work closely with Imams (the local religious leaders) so that they can be engaged in delivering spiritual support to the patients to address their “spiritual distress.” The team endeavors to fulfill their last wishes, including session with spiritual leaders and arranging burial rituals and wishes. [3].

**Rehabilitative care:** To maximize the mobility, functionality, autonomy, and comfort of patients, rehabilitative care is integrated into PALLI-PHC. Physiotherapists work as part of the palliative care team to conduct physiotherapy at the facility and household level. Occupational therapies are also provided along with adaptation, exercises, and assistive devices (e.g. crutches, walkers, commode chairs, kneecaps, wheelchairs) to improve day-to-day activities [3].

**End of life care:** PALLI-PHC offers a comprehensive End of Life Care (EOL) package to ensure comfort, dignity, and compassion during a person’s ending phase of life [3]. This includes timely identification of physical and psychological symptoms specific to this phase of life focusing on optimal symptom control without any distressful clinical interventions, and bereavement support. PALLI-PHC emphasizes respecting individual preferences, including documenting last wishes, preference of place of care, and death [3].

**HEALTH SYSTEM SUPPORT CHAIN**

The whole service delivery in PALLI-PHC is supported through a health system support chain comprising multi-disciplinary engagement, health workforce, advocacy and capacity building, financing, community engagement, supply of essential medical logistics, and information management. Many of the components in the support chain are aligned with WHO health system building blocks [20].

**Health Workforce:** The PALLI-PHC brings a cost-effective solution for efficiently using the health workforce in a crisis setting. The model provides palliative care at nine health facilities and home-based care at seven camps and two host community localities with a force of 23 healthcare workers. This force includes four medical doctors, six nurses, five medical assistants, seven palliative care assistants and two physiotherapists. The staff is divided into multiple teams to deliver palliative care on a rotation basis. Additionally, general medical staff of the health facilities are trained to deliver basic palliative care services during non-clinic hours. The programme also engages frontline palliative care assistants to work closely with the network of Community Health Workers (CHWs) to identify and refer eligible patients in the community.

**Multidisciplinary collaboration:** The multidisciplinary approach in palliative care ensures holistic support for the patients and their family members with consideration of their physical, mental, psychological, social, and spiritual needs [21]. The palliative care team engages the local stakeholders, including the camp-in-charge, site management, protection, and WASH sector actors, to link the patient and the family with social support. Close coordination and referral systems are maintained at PHC/hospital level to ensure clinical care.

**Community Engagement:** Community engagement empowers patients, families and the community, and promotes trust and sustainability of the care model. The PALLI-PHC model promotes engagement of the community health volunteers and religious leaders (e.g., Imams), builds communication with local community representatives, promotes home-based care, mobilize community-based support, and plans intervention-responsive to local culture and rituals (e.g., religious customs, spiritual beliefs, integrative medicine). Palliative care remains integrated with IOM's complaint and feedback mechanisms and community health facility group (CHFG) interventions.

**Capacity building:** Due to the unavailability of palliative care specialists in low-cost settings and considering palliative care is still a new theme for healthcare workers; the PALLI-PHC model deliberately emphasizes extensive capacity building of healthcare workers. Formal training for palliative care and regular primary healthcare team members are organized in collaboration with specialized institutions. Other capacity building activities include supporting supervision, health facility-based CMEs, annual review workshops and weekly meetings, and production of education materials. The programme also organizes orientation sessions with spiritual leaders and community health volunteers.

**Advocacy:** PALLI-PHC encourages advocacy with a wide range of stakeholders for greater integration of palliative care into essential health service packages. This includes sharing the best practices in health sector, celebration of World Hospice and Palliative Care Day, sensitization meetings with community leaders, and social advocacy workshops with local government and service providers. IOM successfully integrated palliative care at two Upazila Health Complexes – the sub-district level government hospitals.

**Monitoring and evaluation:** To ensure the quality and effectiveness of the service, monitoring and evaluation remain a crucial component of the health system support chain. A kobo-based data collection tool is used for recording clinical data, which are analyzed periodically to access the coverage, accessibility and pain, disability and symptom control outcomes. FAMCARE-P16, a self-report scale, is used to assess client satisfaction [22].

Essential medicine in palliative care: To ensure constant availability of medicines and consumables, IOM integrated a list of essential medicines into its supply chain, adapted from WHO Essential Medicines, Essential Medicines for Palliative Care in MSF, and national guideline [14, 23].

**Health Financing:**  While PALLI-PHC itself a cost-efficient model and requires a minimal investment of financial resources, IOM ensures that all needs are addressed in financial planning, including programme budgeting, national and global appeals, and proposals to humanitarian donors.
